# Localization and removal of nonpalpable contraceptive implants: experience from a teaching hospital in Ethiopia

**DOI:** 10.1101/2022.05.10.22274920

**Authors:** Ferid A. Abubeker, Tesfaye H. Tufa, Lemi Belay, Abraham Fessehaye Sium, Jaclyn M Grentzer, Mekdes Bahru Welderufael, Sarah Prager

## Abstract

**Background:** Contraceptive implants are highly effective long-acting reversible contraceptive methods with less than 1 pregnancy per 100 women over the duration of use. In most circumstances, insertion and removal of contraceptive implants is a short and safe procedure performed in an ambulatory setting. Nonpalpable implants are a rare occurrence observed in about 1 per 1,000 insertions but pose a significant challenge. Different management strategies for nonpalpable implants have been described. This report aims to present our approach and experience with localization and removal of nonpalpable implants.

**Methods:** We conducted a facility-based retrospective review of patients evaluated for a nonpalpable contraceptive implant between September 2019 and March 2022 at St. Paul’s Hospital Millennium Medical College (SPHMMC) located in Addis Ababa, Ethiopia. It is a tertiary teaching hospital with Obstetrics and Gynecology (OBGYN) residency as well as a Family Planning fellowship program.

**Results:** Of the 68 cases reviewed close to one-third were referred from other health care facilities with the diagnosis of a nonpalpable implant. Twenty-four (35.3%) patients had at least one previous failed attempt at removal before referral. On ultrasound examination, 27 (40.3%) implants were found below the muscle fascia. Implant removal procedures were successfully done at the outpatient clinic in 65 (95.6%) cases including 40/40 (100%) suprafascial and 25/27 (92.6%) subfascial implants. All removals were performed by OBGYNs with subspecialty training in Family Planning (graduates of a family planning fellowship) or current fellows supervised by subspecialists. In all cases managed at the center, no post-procedure complications have been documented.

**Conclusion:** Our findings show that meticulous evaluation and careful patient selection are essential for the successful localization and removal of nonpalpable implants in outpatient settings. Blind removal attempts prior to appropriate localization should be avoided and referral pathways to specialty centers should be arranged.

## Introduction

Contraceptive implants are highly effective long-acting reversible contraceptive methods with less than 1 pregnancy per 100 women over the duration of use (1, 2). Globally more than 20 million women are using contraceptive implants (3). Over the past decade, there has been a significant increase in the uptake of contraceptive implants in sub-Saharan African counties (4, 5). In Ethiopia, the use of contraceptive implants has increased from 2.3% in 2011 to 9% in 2019 (6, 7).

The national family planning program of Ethiopia offers contraceptive implants free of charge at different levels of health care facilities including primary care units. In addition, insertion and removal of contraceptive implants are provided by a widely ranging cadre of trained health care providers. Contraceptive implant devices provided by the national program include Jadelle® (two rods, each containing 75 mg of levonorgestrel) and Nexplanon® (a newer radiopaque version of the implant Implanon®, with one rod containing 68 mg of etonogestrel) (8).

The provision of contraceptive implants requires comprehensive training on insertion and removal techniques to minimize complications (9). In most circumstances, insertion of contraceptive implants is a short and safe procedure performed in an ambulatory setting (10). However, very rarely, serious complications such as deep insertion, migration, and intravascular insertion with pulmonary embolization have been reported (9, 11, 12).

To minimize insertion errors and improve localization, the manufacturer of the etonogestrel implant made modifications to the applicator and added 15 mg of barium sulfate to the implant device making it radiopaque and thus visible on x-ray (13). They also changed the recommended insertion site from the sulcus between the biceps and triceps muscles to the site immediately overlying the triceps muscle about 8-10 cm from the medial epicondyle of the humerus and 3-5 cm posterior to the sulcus. This insertion site change was intended to minimize the risk of injury to neurovascular structures within and around the sulcus (14).

When the implant is palpable, removal is usually easy, quick, and can be done without imaging guidance (10, 15). Nonpalpable implants are encountered uncommonly, in about 1 per 1,000 insertions (9, 16). However, the management of nonpalpable implants poses a significant challenge. Studies report difficulty in localizing nonpalpable implants and complications from iatrogenic neurovascular injury during removal attempts (17, 18).

Different strategies were instituted to localize and remove nonpalpable implants. Some health care providers implemented intraoperative fluoroscopy or referral to an orthopedic surgeon (19) or peripheral nerve surgeon (20). Others introduced ultrasound localization followed by removal using a modified vasectomy clamp (21), or removal under continuous ultrasound guidance (22). Some settings also described the establishment of specialty referral centers to manage cases of nonpalpable implants by specifically trained care providers (23, 24).

Our clinical impression is that over the past several years, the rate of nonpalpable or missed implants has markedly increased. However, to the best knowledge of the authors, there are no documented reports about this complication in Ethiopia. This report aims to present our approach and experience with localization and removal of nonpalpable implants. By compiling this summary, we hope to help care providers improve the clinical management of patients. We also believe the information will enable care providers, program managers, and policymakers to develop strategies to minimize this complication and optimize outcomes.

## Materials and methods

### Setting

We conducted a facility-based retrospective review of patients evaluated for a nonpalpable contraceptive implant between September 2019 and March 2022 at St. Paul’s Hospital Millennium Medical College (SPHMMC), located in Addis Ababa, Ethiopia. It is a tertiary teaching hospital with Obstetrics and Gynecology (OBGYN) residency as well as a Family Planning fellowship program. Contraceptive services are provided in a dedicated reproductive health clinic called MICHU clinic. The clinic receives referrals from all regions of the country for complex contraception care including removal of nonpalpable contraceptive implants.

### Client approach

Clients who presented to the family planning clinic for implant removal were first evaluated by an OBGYN resident or midwife/nurse working at the clinic. Those with nonpalpable implants were then referred to the Family Planning division.

We began our evaluation with thorough palpation of the entire upper arm starting from the level of the epicondyles to the axilla in a systematic fashion. For those patients known to have a radiopaque implant (either Nexplanon® or Jadelle®), a two-view X-ray of the upper arm was performed to confirm the presence of the device.

In all cases, ultrasound localization of the implant was done with mapping on the skin surface using surgical markers. Ultrasound localization was performed at the family planning clinic using a high-frequency, large footprint, linear array transducer (Samsung SonoAce R7, 12-15 MHz, LN5-12 system). During imaging, patients are positioned supine on the examination table with arms flexed 90° at the elbow and externally rotated exposing the medial aspect of the upper arm.

Scanning was started from the site where the implant was presumed to be present based either on skin findings from previous insertion or removal attempts (scars) or X-ray findings. Attempts were made to first visualize the implant with the probe in the transverse plane where we would expect to see an echogenic spot with the characteristic posterior acoustic shadow. We also assessed the location of the implant device relative to the muscle fascia and employed color Doppler to evaluate the proximity to vascular structures. The probe was then turned 90 degrees to obtain a longitudinal view of the implant and measure its length.

For implants not detected on ultrasound, we obtained an X-ray of the upper arm, if not previously done. In cases where neither upper arm ultrasound nor X-ray locates the implant, two view chest X-ray was performed. MRI of the upper arm or chest was obtained for implant localization in patients with a known Implanon®, as it is not radiopaque.

### Removal procedure

After localization of the implant, device removal was performed under local anesthesia in the outpatient setting. In cases where the implant was found very close to vascular structures, removal was scheduled in the operating room with an orthopedic or general surgeon consultation.

The proposed incision site was prepared with an antiseptic solution and covered with a fenestrated drape. After administering 1-2ml of 1% lidocaine, a 5mm longitudinal incision was made. For suprafascial implants, we bluntly dissected the subcutaneous tissue with mosquito forceps. For subfascial implants, blunt dissection of the subcutaneous tissue continued to the level of the fascia. The fascia was then grasped with two forceps and opened longitudinally by sharp dissection. If unable to localize the implant during dissection, the surgeon localized the implant by palpating with a finger through the incision or ultrasound guidance as needed. The implant device was then grasped with mosquito and brought to the skin surface. Any tissue capsule around the device was released by blunt or sharp dissection. The implant was then removed and ensured to be intact and completely removed. The incision was closed in 1 or 2 layers using absorbable sutures. Patients were provided with oral analgesics and instructed to return for any complications.

### Study data collection and analysis

We reviewed medical charts of patients who were evaluated for nonpalpable contraceptive implants. Demographic characteristics, implant-related data, clinical evaluation, and outcomes of patients were collected using a structured questionnaire template prepared on an android platform. Collected data was sent to a central server and then exported to and analyzed using IBM

SPSS Statistics version 25.0 for Windows (IBM Corp., Armonk, NY, USA). Descriptive analysis is used to present the findings of this study.

### Ethical considerations

Prior to data collection, the study proposal was submitted to the Institutional Review Board (IRB) of SPHMMC for ethical review and approval. Sequential numbers were assigned to cases as data collection was conducted to ensure confidentiality. No names of patients or managing health professionals were included in the study.

## Results

A total of 77 medical records were identified of which 9 cases were excluded due to incomplete data. Thus, a total of 68 cases were reviewed in this study. The mean age was 27.2±4.9 years. Close to one-third of patients were referred from other health care facilities with the diagnosis of nonpalpable implant. More than 90% of the patients had etonogestrel implants and interval insertions accounted for 54.4% of the cases. Twenty-four (35.3%) patients had at least one previous failed attempt at removal before referral. (Table 1).

**Table 1:** Demographic and implant characteristics of patients presenting with nonpalpable contraceptive implants.

| Characteristics | Frequency (n) | Percentage (%) |
| --- | --- | --- |
| **Age (years)** |  |  |
| < 25 | 23 | 33.8 |
| 25-34 | 38 | 55.9 |
| ≥ 35 | 7 | 10.3 |
| **Parity** |  |  |
| 0 | 17 | 25.0 |
| 1 | 26 | 38.2 |
| 2 | 14 | 20.6 |
| ≥ 3 | 11 | 16.2 |
| **Patient living address** |  |  |
| Within Addis Ababa | 37 | 54.4 |
| Outside Addis Ababa | 31 | 45.6 |
| **Source of referral** |  |  |
| From SPHMMC/Internal | 7 | 10.3 |
| Self-referral | 13 | 19.1 |
| Health facility outside Addis Ababa | 22 | 32.4 |
| Health facility within Addis Ababa | 26 | 38.2 |
| **Reason for referral from facilities** |  |  |
| Nonpalpable implant | 17 | 35.5 |
| Deeply inserted implant | 10 | 20.8 |
| Missed implant | 10 | 20.8 |
| Failed removal attempt | 10 | 20.8 |
| Possible migration | 1 | 2.1 |
| **Type of implant device** |  |  |
| Nexplanon® | 52 | 76.5 |
| Implanon® | 11 | 16.2 |
| Jadelle® | 5 | 7.3 |
| **Timing of implant insertion** |  |  |
| Interval | 37 | 54.4 |
| Postpartum | 19 | 28.0 |
| Postabortion | 7 | 10.3 |
| Undocumented | 5 | 7.3 |
| **Reason for implant removal** |  |  |
| On recommended time | 30 | 44.1 |
| Desire for pregnancy | 26 | 38.2 |
| Side effects | 12 | 17.7 |
| **Removal attempt before referral** |  |  |
| 0 | 44 | 64.7 |
| 1 | 18 | 26.5 |
| ≥2 | 6 | 8.8 |

In 18 (26.5%) patients, X-ray of the upper arm followed by an ultrasound examination was used to localize the implant device before successful removal. For 50 (73.5%) cases, point of care ultrasound examination was performed as the initial investigative modality at the family planning clinic. All but two cases had a successful removal procedure on the first attempt at the clinic. The two patents with a failed initial removal attempt had the older Implanon® device and MRI was used for better localization. Repeat ultrasound examination was also done to identify the typical posterior acoustic shadow and aid the identification of the incision site. The subsequent removal procedure was successful in both cases. (Fig 1).

**Fig 1.**
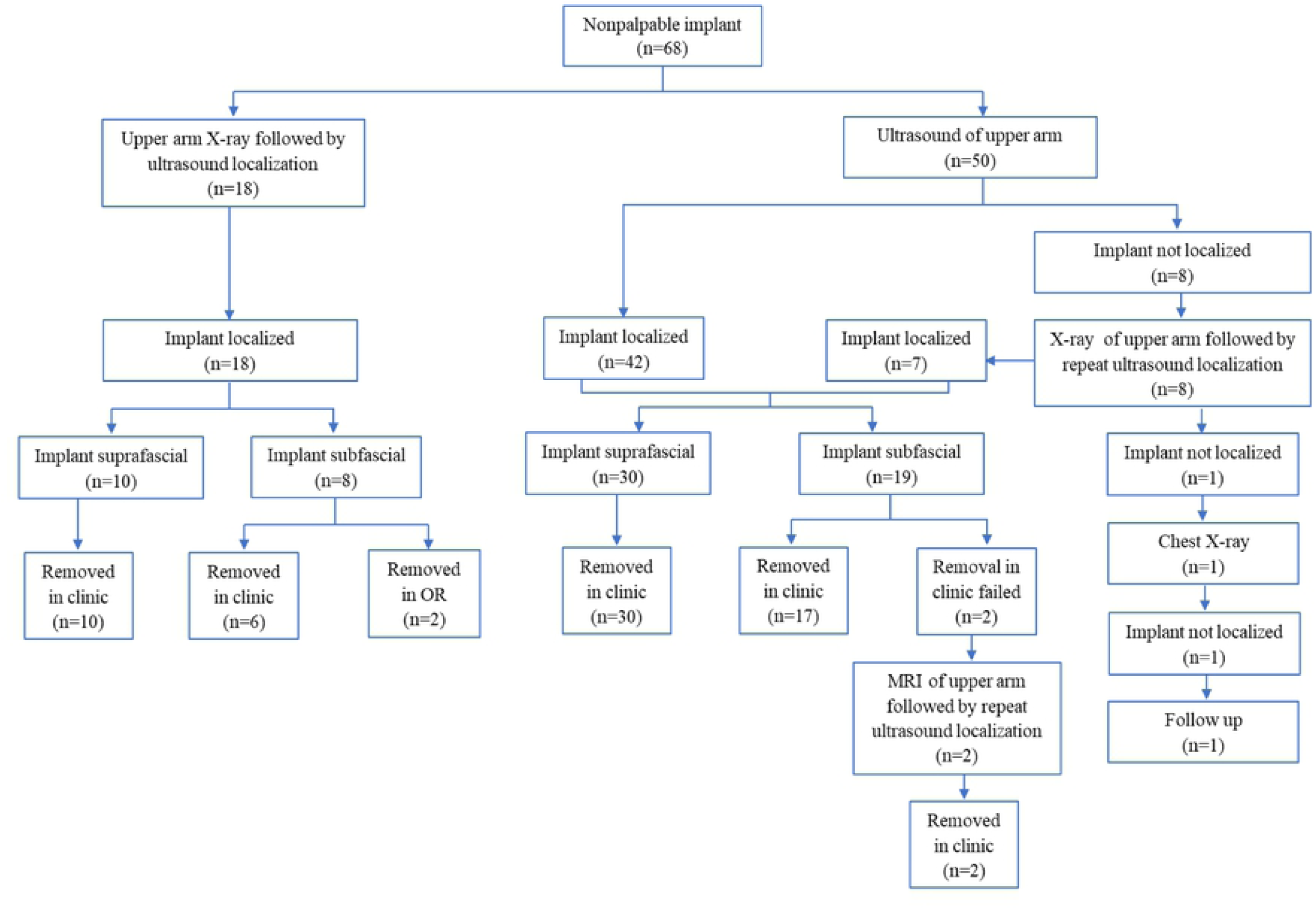
Flow chart demonstrating the clinical evaluation and management outcome of patients presenting with nonpalpable contraceptive implants.

On ultrasound examination, 27 (40.3%) implants were found below the muscle fascia. Fifty-five patients (82.0%) had documentation of the depth of the implant from the skin surface. Among these, the mean depth of the implant was 5.9 ± 2.6 mm below the skin.

Implant removal procedures were successfully done at the outpatient clinic in 65 (95.6%) cases including 40/40 (100%) suprafascial and 25/27 (92.6%) subfascial implants. In two cases, subfascial implants were removed in the operating room due to the close proximity of the implant to vascular structures on color Doppler study. (Fig 1).

One patient presented for removal of Nexplanon device that was inserted three years prior on the first postpartum day. We failed to localize the device with ultrasound and X-ray imaging of the upper arm. With the suspicion of possible migration, chest X-ray was done but the findings were unremarkable. She could not afford other imaging studies such as CT-Scan and MRI, and determination of serum etonogestrel is not available in this setting. Though she initially had an intention to switch to injectable contraceptive, she later changed her mind and declined to use any form of contraceptive method. Currently, she is on follow-up.

All removals were performed by OBGYNs who completed subspecialty training in a Family Planning fellowship, or by Family Planning fellows supervised by subspecialists. For the two cases that underwent removal in the operating room, preoperative discussion and preparations were made with an experienced surgeon for possible intraoperative consultation should the need arise. In all cases managed at SPHMMC, no post-procedure complications have been documented.

## Discussion

Contraceptive implants are among the most effective contraceptive methods (25). Recent figures show an increasing trend in the utilization of implants across different settings (3, 4). With the increased uptake, the incidence of uncommon complications such as deep insertions and difficult removals will likely increase (26).

Meticulous insertion techniques have been shown to decrease complications encountered during removal procedures (15). Indeed, the manufacturer of Nexplanon® has recently updated the instructions for the insertion of the device (14). However, it is worth noting that the Ethiopian national family planning training manual is lagging behind and has not been updated to reflect this most recent guidance. All the patients in this study with an etonogestrel implant had the device inserted based on the older insertion site recommendation in the sulcus between the biceps and triceps muscles.

In rare cases where the implant is not easily palpable, appropriate clinical and imaging evaluation should be performed to confirm the presence and location of the device before any removal attempt is made (27, 28). In our study more than one-third of patients had at least one previous failed attempt of removal before referral. It is of paramount importance that providers avoid blind removal procedures. Serious iatrogenic neurovascular injuries have been reported following removal procedures before the appropriate localization of the device (18, 29, 30). Fortunately, no patient presented to our clinic with procedure-related complications.

Almost all patients presenting to our clinic for removal of nonpalpable implants were successfully managed in the outpatient setting. We evaluated each patient carefully and performed thorough preoperative planning to identify cases that required surgical consultation for an interdisciplinary approach. Our findings are in line with the experience of specialty centers in the US (23, 26), the UK (31), and South Africa (24). These studies showed that removal of nonpalpable implants is best performed by specially trained practitioners. Thus, referrals to family planning specialists with experience managing difficult implant removals should be considered. Our center is currently the only family planning specialty center in the nation with a dedicated family planning fellowship program. To our knowledge, there is only one other facility in sub-Saharan Africa providing specialty care for nonpalpable and difficult implant removals (32).

Our approach to patients presenting with nonpalpable implants has evolved through time. For some of the patients, our initial practice involved X-ray imaging of the upper arm to confirm the presence of the device followed by ultrasound localization before the removal procedure. We recently updated our algorithm and currently perform high-resolution ultrasound examinations at the family planning clinic as the initial investigative modality. If the initial point of care ultrasound examination fails to localize the device, upper arm X-ray imaging is done to confirm the presence of the device and guide subsequent ultrasound localization. (Fig 2).

**Fig 2.**
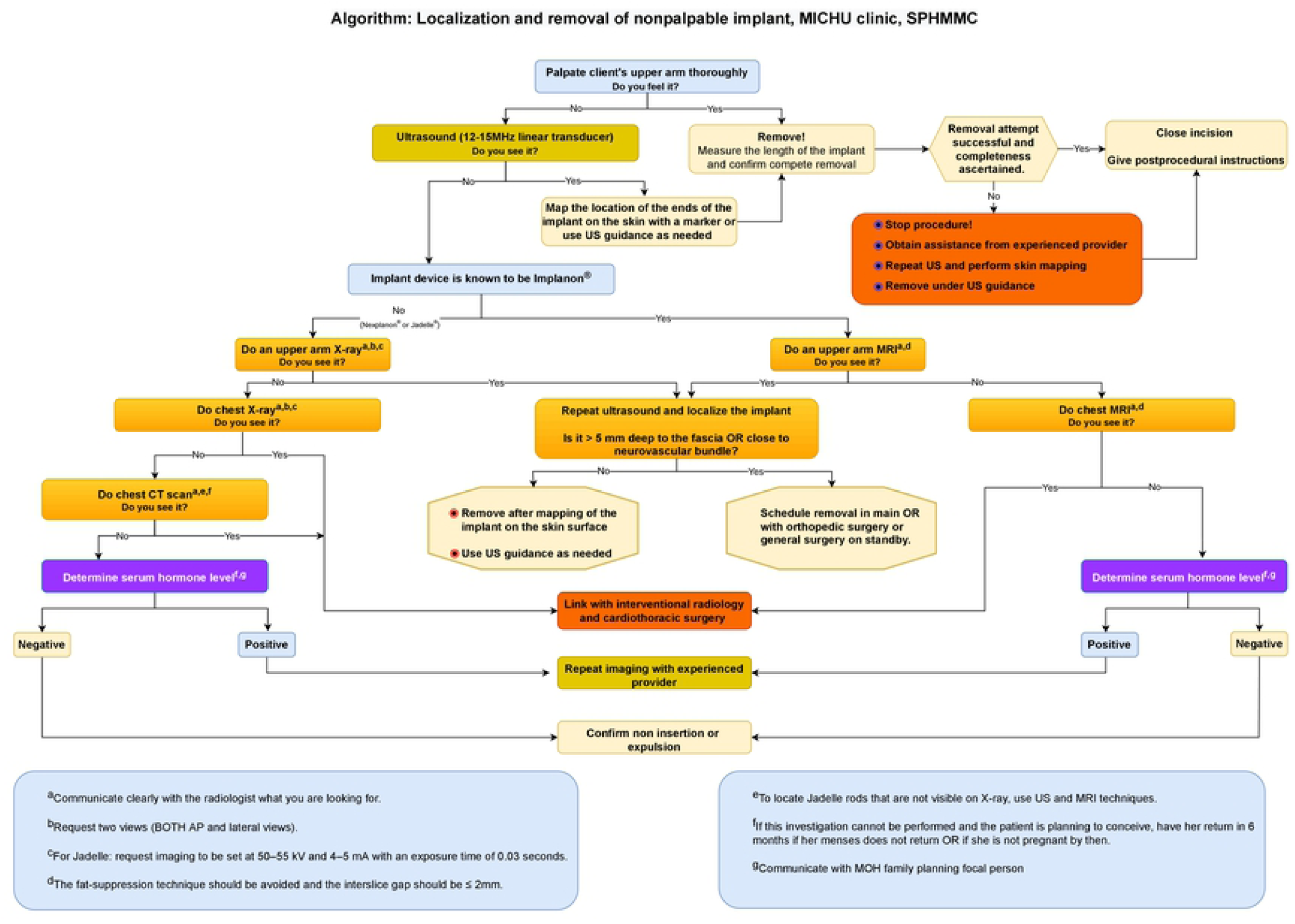
Algorithm for localization and removal of nonpalpable contraceptive implants.

This approach avoids delays from waiting for X-ray reports and typically allows for same-day implant localization and removal. This is highly relevant in our setting where nearly half of patients coming to the facility are living outside of Addis Ababa. Others have also reported the importance of incorporating point of care ultrasound localization in their practice (24, 26).

Once a nonpalpable implant is localized several removal techniques have been described(19-22). The utilization of a modified vasectomy clamp with a 2.2 mm ring diameter has gained more popularity as it prevents crushing and fracture of the implant device(21, 23, 32, 33). Due to the lack of this instrument, we routinely use mosquito forceps to carefully remove the implant.

## Conclusion

Our findings show that meticulous evaluation and careful patient selection are essential for the successful localization and removal of nonpalpable implants in outpatient settings. Providers should be familiar with the workup of patients presenting with nonpalpable implants. Blind removal attempts prior to appropriate localization should be avoided and referral pathways to specialty centers should be arranged. In addition, these specialty centers need to have the essential medical equipment and trained personnel to deliver and maintain high-quality services.

## Data Availability

All relevant data are within the manuscript and its Supporting Information files.

## Acknowledgments

We would like to express our deepest gratitude to the department of OBGYN at SPHMMC and the staff working at MICHU clinic.

## References

1. World Health Organization Department of Reproductive Health and Research (WHO/RHR) and Johns Hopkins Bloomberg School of Public Health/Center for Communication Programs (CCP) KfHP. Family planning: a global handbook for providers (2018 update). Baltimore and Geneva: CCP and WHO. 2018.

2. Power J, French R, Cowan FM. Subdermal implantable contraceptives versus other forms of reversible contraceptives or other implants as effective methods for preventing pregnancy. Cochrane Database of Systematic Reviews. 2007(3).

3. United Nations, Department of Economic and Social Affairs, Population Division (2019). Contraceptive Use by Method 2019 Data Booklet.

4. Jacobstein R. Liftoff: the blossoming of contraceptive implant use in Africa. Global Health: Science and Practice. 2018;6(1):17–39.

5. Ahmed S, Choi Y, Rimon JG, Alzouma S, Gichangi P, Guiella G, et al. Trends in contraceptive prevalence rates in sub-Saharan Africa since the 2012 London Summit on Family Planning: results from repeated cross-sectional surveys. The Lancet Global Health. 2019;7(7):e904–e11.

6. Central Statistical Agency [Ethiopia] and ICF. Ethiopia Demographic and Health Survey 2011. Addis Ababa, Ethiopia, and Calverton, Maryland, USA: Central Statistical Agency and ICF International. 2012.

7. Ethiopian Public Health Institute (EPHI) [Ethiopia] and ICF. Ethiopia Mini Demographic and Health Survey 2019: Key Indicators. Rockville, Maryland, USA: EPHI and ICF. 2019.

8. Ministry of Health, Federal Democratic Republic of Ethiopia. National Guideline for family planning services in Ethiopia. Addis Ababa: Ethiopia: FMOH; 2020.

9. Creinin MD, Kaunitz AM, Darney PD, Schwartz L, Hampton T, Gordon K, et al. The US etonogestrel implant mandatory clinical training and active monitoring programs: 6-year experience. Contraception. 2017;95(2):205–10.

10. Blumenthal PD, Gemzell-Danielsson K, Marintcheva-Petrova M. Tolerability and clinical safety of Implanon(r). The European Journal of Contraception & Reproductive Health Care. 2008;13(sup1):29–36.

11. Rivera F, Bianciotto A. Contraceptive subcutaneous device migration: what does an orthopaedic surgeon need to know? A case report and literature review. Acta Bio Medica: Atenei Parmensis. 2020;91(4-S):232.

12. Rowlands S, Mansour D, Walling M. Intravascular migration of contraceptive implants: two more cases. Contraception. 2017;95(2):211–4.

13. Mansour D. Nexplanon(r): what Implanon(r) did next. The Journal of Family Planning and Reproductive Health Care. 2010;36(4):187.

14. Merck Sharp & Dohme Limited. Direct Healthcare Professional Communication on the association of Nexplanon-etonogestrel 68mg, implant for subdermal use - Update to the insertion and removal instructions to minimise the risks of neurovascular injury and implant migration. 15 January 2020.

15. Shulman LP, Gabriel H. Management and localization strategies for the nonpalpable Implanon rod. Contraception. 2006;73(4):325–30.

16. Reed S, Do Minh T, Lange JA, Koro C, Fox M, Heinemann K. Real world data on Nexplanon(r) procedure-related events: final results from the Nexplanon Observational Risk Assessment study (NORA). Contraception. 2019;100(1):31–6.

17. Restrepo CE, Spinner RJ. Major nerve injury after contraceptive implant removal: case illustration. J Neurosurg. 2016;124(1):188–9.

18. Lefebvre R, Hom M, Leland H, Stevanovic M. Peripheral nerve injury with Nexplanon removal: case report and review of the literature. Contraception and Reproductive Medicine. 2018;3(1):1–6.

19. Hellwinkel JE, Konigsberg MW, Oviedo J, Castaño PM, Kadiyala RK. Subfascial-located contraceptive devices requiring surgical removal. Contraception and Reproductive Medicine. 2021;6(1):1–4.

20. Odom EB, Eisenberg DL, Fox IK. Difficult removal of subdermal contraceptive implants: a multidisciplinary approach involving a peripheral nerve expert. Contraception. 2017;96(2):89–95.

21. Chen MJ, Creinin MD. Removal of a nonpalpable etonogestrel implant with preprocedure ultrasonography and modified vasectomy clamp. Obstet Gynecol. 2015;126(5):935–8.

22. Jacques T, Brienne C, Henry S, Baffet H, Giraudet G, Demondion X, et al. Minimally invasive removal of deep contraceptive implants under continuous ultrasound guidance is effective, quick, and safe. Eur Radiol. 2022;32(3):1718–25.

23. Matulich MC, Chen MJ, Schimmoeller NR, Hsia JK, Uhm S, Wilson MD, et al. Referral center experience with nonpalpable contraceptive implant removals. Obstet Gynecol. 2019;134(4):801.

24. Petro G, Spence T, Patel M, Gertz AM, Morroni C. Difficult etonogestrel implant removals in South Africa: a review of 74 referred cases. Contraception. 2020;102(2):129–32.

25. Darney P, Patel A, Rosen K, Shapiro LS, Kaunitz AM. Safety and efficacy of a single-rod etonogestrel implant (Implanon): results from 11 international clinical trials. Fertil Steril. 2009;91(5):1646–53.

26. Mastey N, Matulich MC, Uhm S, Baker CC, Melo J, Chen MJ, et al. US referral center experience removing nonpalpable and difficult contraceptive implants with in-office ultrasonography: A case series. Contraception. 2021;103(6):428–30.

27. Kim S, Choi YS, Kim JS, Kim S, Cho S. Experiences of localization and removal of non-palpable subdermal contraceptive implants with ultrasound. Obstetrics & Gynecology Science. 2019;62(3):166–72.

28. Mansour D, Fraser IS, Walling M, Glenn D, Graesslin O, Egarter C, et al. Methods of accurate localisation of non-palpable subdermal contraceptive implants. J Fam Plann Reprod Health Care. 2008;34(1):9.

29. Monteiro RB, Metzger PB, Moura ABd, Silva Filho AH, Campos MN, Brito ASd, et al. Traumatic pseudoaneurysm in brachial artery after removal of a subdermal contraceptive implant. Jornal Vascular Brasileiro. 2020;19.

30. Laumonerie P, Blasco L, Tibbo ME, Leclair O, Kerezoudis P, Chantalat E, et al. Peripheral nerve injury associated with a subdermal contraceptive implant: illustrative cases and systematic review of literature. World Neurosurg. 2018;111:317–25.

31. Mansour D. UK provision for removal of non-palpable contraceptive implants. J Fam Plann Reprod Health Care. 2009;35(1):3–4.

32. Petro G. Non-palpable and difficult contraceptive implant removals: The New Somerset Hospital referral-clinic experience. S Afr J Obstet Gynaecol. 2017;23(3):101–4.

33. Mansour D, Walling M, Glenn D, Egarter C, Graesslin O, Herbst J, et al. Removal of non-palpable etonogestrel implants. The Journal of Family Planning and Reproductive Health Care. 2008;34(2):89.

